# NOVEL RT-qPCR ASSAYS ENABLE RAPID DETECTION AND DIFFERENTIATION BETWEEN SARS-COV-2 OMICRON (BA.1) AND BA.2 VARIANTS

**DOI:** 10.1101/2022.02.22.22271222

**Authors:** Oran Erster, Areej Kabat, Hadar Asraf, Virginia Levy, Batya Mannasse, Roberto Azar, Ital Nemet, Limor Kliker, Shay Fleishon, Michal Mandelboim, Ella Mendelson, Neta S Zuckerman

**Affiliations:** Central Virology Laboratory, Public Health Services, Ministry of Health, Chaim Sheba Medical Center, Ramat Gan, Israel; Israel Institute for Biological Research; School of Public Health, Sackler Faculty of Medicine, Tel-Aviv University, Tel-Aviv, Israel

**Keywords:** SARS-COV2, Omicron, variants, BA.1, BA.2, Molecular Diagnostics, qPCR, differentiation

## Abstract

In this report, we describe the development and initial validation of novel SARS-COV-2 Omicron-specific reactions that enable the identification of Omicron (BA.1) and BA.2 variants. Mutations that are either shared by both BA.1 and BA.2, or are exclusive for BA.1 or for BA.2 were identified by bioinformatic analysis, and corresponding probe-based quantitative PCR reactions were developed to identify them. We show that multiplex combinations of these reactions provide a single-reaction identification of the sample as BA.1, BA.2, or as non-Omicron SARS-COV-2. All four reactions described herein have a sensitivity of less than ten copies per reaction, and are amendable for multiplexing. The results of this study suggest that the new assays may be useful for testing both clinical and environmental samples to differentiate between these two variants.

## INTRODUCTION

The emergence of the SARS-COV-2 Omicron variant, formerly designated B.1.529, and currently designated BA.1, resulted in resurgence of morbidity and increased mortality worldwide, including countries with high vaccination rates, where the SARS-COV-2 Delta variant circulation already decreased (Elliott et al. 2022). Several weeks after the global spreading of the BA.1 variant, a subsequent spreading of a related variant, BA.2, was observed in several countries (Desingu and Nagarajan, 2022). The global circulation of BA.2 increases gradually, in parallel with the on-going circulation of BA.1. The epidemiological and medical consequences of the co-circulation these two variants are not well understood, as well as the differences between them, with respect to infectivity, pathogenicity and vaccine evasion (Pulliam et al. 2021, Yu et al. 2022). The rapid pace in which SARS-COV-2 (SC-2) variants emerge and spread globally, poses a substantial challenge for efficient surveillance, rendering cell culturing and whole genome sequencing often insufficient for providing fast and reliable data. We previously demonstrated that rapid, affordable and high throughput differentiation between co-circulating SC-2 variants can be accomplished using multiplex quantitative PCR (qPCR), and that this approach can complement and, in some cases, outperform genomic sequencing and similar approaches, for SC-2 variant identification (Erster et al. 2021a, Erster et al. 2021b). More recently, we reported on the development of qPCR-based assays that specifically identify BA.1 samples and allow straightforward scaling-up for high throughput testing (Erster et al. 2022). In this report, we describe, to our knowledge for the first time, differential RT-qPCR assays that rapidly distinguish between BA.1 and BA.2.

## 2. MATERIALS AND METHODS

### 2.1 Clinical samples

Clinical samples were prepared as described previously (Erster et al. 2021a). Briefly, nasopharyngeal swabs were collected from patients for diagnostic purpose. RNA was extracted and used for clinical testing. Samples were processed and RNA was extracted as described previously (doi.org/10.1101/ 2021.10.11.21264831).

### 2.2. Cell culture

Nasopharyngeal swab samples were used to infect Vero-6 cells as described previously (doi.org/10.1101/ 2021.10.11.21264831). Upon the onset of cytopathic effect (CPE), the medium was collected and cultured virus was examined by RNA extraction and subsequent PCR.

### 2.3. Design of BA.1 and BA.2-specific reactions

Identification of BA.1 and BA.2 specific mutations was performed by aligning complete genomes of BA.1, BA.2, A19 (Wuhan SC-2) and Delta. Genome sequences were obtained from the GISAID initiative website (https://www.gisaid.org/). Sequences were aligned and analyzed using the Geneious software package (https://www.geneious.com/). Selected regions were examined again using the global analysis of the NextStrain website tools (https://nextstrain.org/sars-cov-2/), where each mutation was tested separately, to ensure its uniqueness. Primers and probes were designed and tested using the Primer 3 program (https://bioinfo.ut.ee/primer3-0.4.0/) embedded in the Geneious software.

### 2.4. RT-qPCR assays

Assembly of quantitative RT-PCR (RT-qPCR) was as described before (doi.org/10.1101/ 2021.10.11.21264831 and doi.org/10.1101/2021.12.07.21267293). Briefly, SensiFast master mix (https://www.meridianbioscience.com/lifescience/) was used with the specific primers and probes combination for each reaction. For environmental samples. The primers and probes sequences and additional data are detailed in **Table 1**. The reaction components are detailed in **Table 2**. The cycling conditions for the Omicron reactions were as follows: 16 min. at45°0, 2’20” at 95°C, 45X [4sec at 95°C, 10sec at 55°C, 10sec at 60°C]. Fluorescence was read during the extension step in each cycle.

**Table 1.**
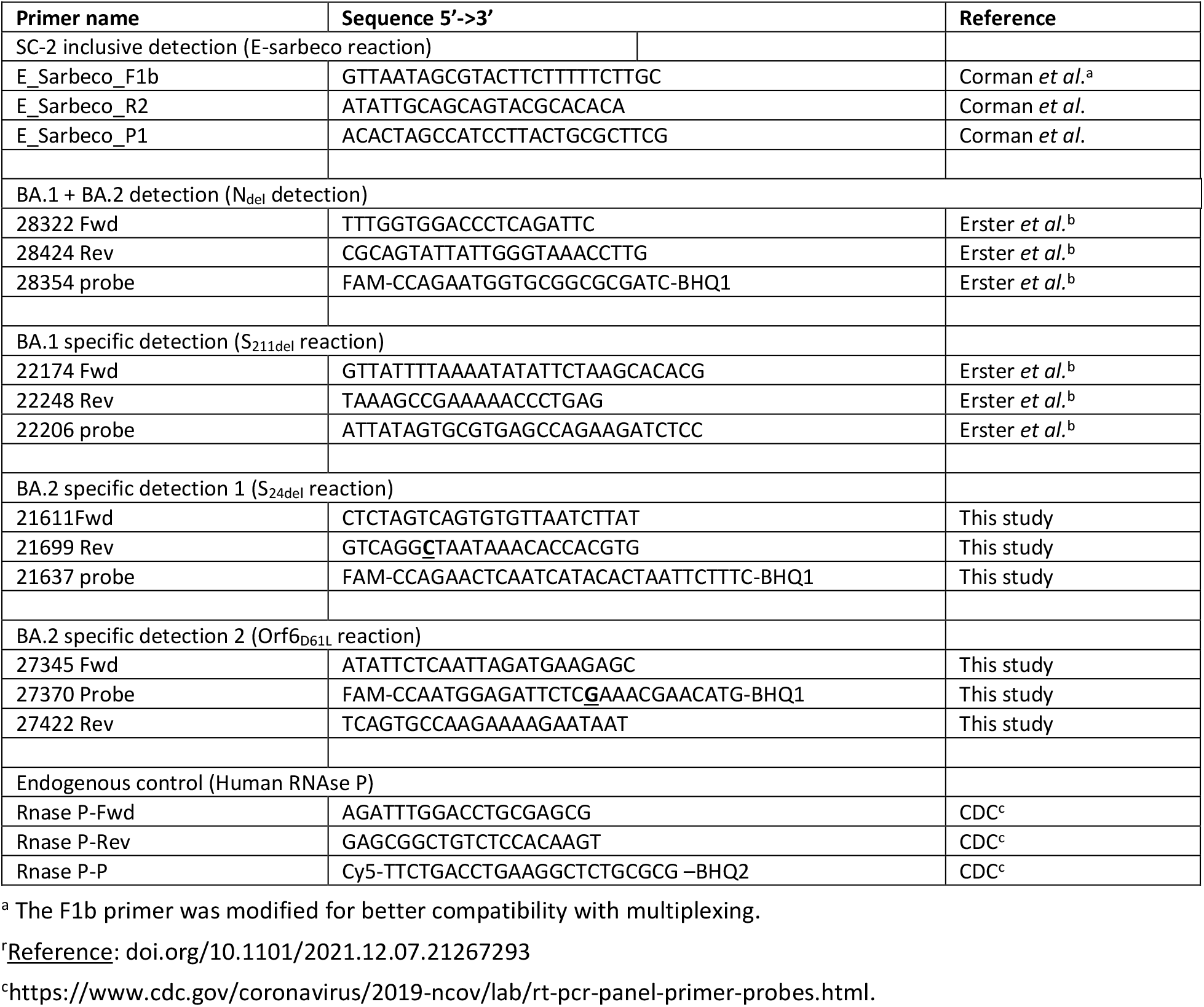
Primers and probes used in this study. Bold, underlined letters represent deliberate mismatch nucleotides that were inserted to increase specificity and minimize secondary structure formation.

**Table 2.** Compositions of the multiplex assays used in this study. The volume (µl) of each component and the stock and final concentrations of the primers and probes are indicated. For each probe, the fluorophore used is indicated. Conc. / Rxn: concentration per reaction. µl/ Rxn: µl per reaction.

| **E-sarbeco + N<sub>31del</sub> + S<sub>24del</sub> + RNaseP** | Conc. / Rxn | μl/ Rxn | **E-sarbeco + Orf6<sub>D61L</sub>** | Conc. / Rxn | μl/ Rxn |
| --- | --- | --- | --- | --- | --- |
| SensiFast Probe One-Step |  | 10 | SensiFast Probe One-step |  | 10 |
| H<sub>2</sub>O |  | 1.55 | H<sub>2</sub>O |  | 3.1 |
| E-Sarbeco-F1b 40μM | 500nM | 0.25 | E-Sarbeco-F1b 40μM | 400nM | 0.2 |
| E-Sarbeco-R 40μM | 500nM | 0.25 | E-Sarbeco-R 40μM | 400nM | 0.2 |
| E-Sarbeco-P TexasRed 20μM | 250nM | 0.25 | E-Sarbeco-P TXred 20μM | 200nM | 0.2 |
| COV19_28322Fwd_F 40uM | 600nM | 0.3 | CoV19 21611Fwd 40uM | 600nM | 0.3 |
| COV19_28412 Rev 40uM | 600nM | 0.3 | Cov19 21699 Rev 40uM | 600nM | 0.3 |
| COV19_28354_P 20uM HEX | 300nM | 0.3 | 21637 Probe FAM 20uM | 300nM | 0.3 |
| CoV19 21611Fwd 40uM | 600nM | 0.3 | RNase inhibitor |  | 0.2 |
| Cov19 21699 Rev 40uM | 600nM | 0.3 | RT-Enzyme SensiFast |  | 0.2 |
| 21637 Probe FAM 20uM | 300nM | 0.3 | RNA |  | 5 |
| RNasP-F | 300nM | 0.15 | **Total** |  | **20** |
| RNasP-R | 300nM | 0.15 |  |  |  |
| RNasP-P / Cy-5 | 200nM | 0.15 |  |  |  |
| RT-Enzyme SensiFast |  | 0.25 | **E-sarbeco + S<sub>211del</sub>** | Conc. / Rxn | μl/ Rxn |
| RNase inhibitor |  | 0.2 | SensiFast Probe One-step |  | 10 |
| RNA |  | 5 | H<sub>2</sub>O |  | 3.1 |
| **Total** |  | **20** | E-Sarbeco-F1b 40μM | 400nM | 0.2 |
|  |  |  | E-Sarbeco-R 40μM | 400nM | 0.2 |
|  |  |  | E-Sarbeco-P TXred 20μM | 200nM | 0.2 |
|  |  |  | COV19_22174Fwd_F 40uM | 600nM | 0.3 |
|  |  |  | COV19_22248 Rev 40uM | 600nM | 0.3 |
|  |  |  | COV19_22206_P 20uM FAM | 300nM | 0.3 |
|  |  |  | RNase inhibitor |  | 0.2 |
|  |  |  | RT-Enzyme SensiFast |  | 0.2 |
|  |  |  | RNA |  | 5 |
|  |  |  | **Total** |  | **20** |

### 2.5 Design and synthesis of In vitro transcribed control standards

In order to perform absolute quantification of target copies using the novel reactions, and to evaluate their analytical sensitivity, RNA targets for each reaction were synthesized, as described previously (doi.org/10.1101/2021.05.19.21257439). Briefly, a region spanning the qPCR target sequence was amplified and transcribed to RNA using an *In vitro* transcription kit (https://www.thermofisher.com/). The resulting *In vitro* transcribed (IVT) RNA was purified, quantified and diluted. Calibration curves were generated for each reaction using the synthesized standards, and the analytical limit of detection was determined.

## ETHICAL STATEMENT

This study was conducted according to the guidelines of the Declaration of Helsinki, and approved by the Institutional Review Board of the Sheba Medical Center institutional review board (7045-20-SMC). Patient consent was waived as the study used remains of clinical samples and the analysis used anonymous clinical data.

## 3. RESULTS

### 3.1. Design of BA.1/BA.2 inclusive and selective PCR tests

In order to allow both inclusive and selective identification of BA.1 and BA.2, complete genome sequences of A19 (Wuhan strain), Delta (B.1.617.2), BA.1 and BA.2 were analyzed, to identify sequences that are unique and common for both BA.1 and BA.2, and sequences that are exclusively unique to either one of them, and are absent from A19 and Delta genomes. Four such regions were identified and selected. The deletion in the Nucleocapsid (N) gene, at amino acid position 31 (N_31del_) is common to both BA.1 and BA.2 (**Figure 1A**). In this reaction, the probe is complement to the deletion site and cannot bind to the WT sequence (**Figure 2A**). The deletion at position 211 and insertion at positions 214-216 of the spike (S) gene are exclusive for variant BA.1 (**Figure 1B, Figure 2B**). The deletion at position 24 of the spike gene and the codon substation at position 61 of the Orf6 gene, are both unique for BA.2 (Orf6_D61L_ and S_24del_, **Figure 1C,D** and **Figure 2C,D**, respectively). Based on these mutations, four probe-based qPCR reactions were designed, where the probe in each reaction is complementary to the mutated sequence, as depicted in **Figure 2**.

**Figure 1.**
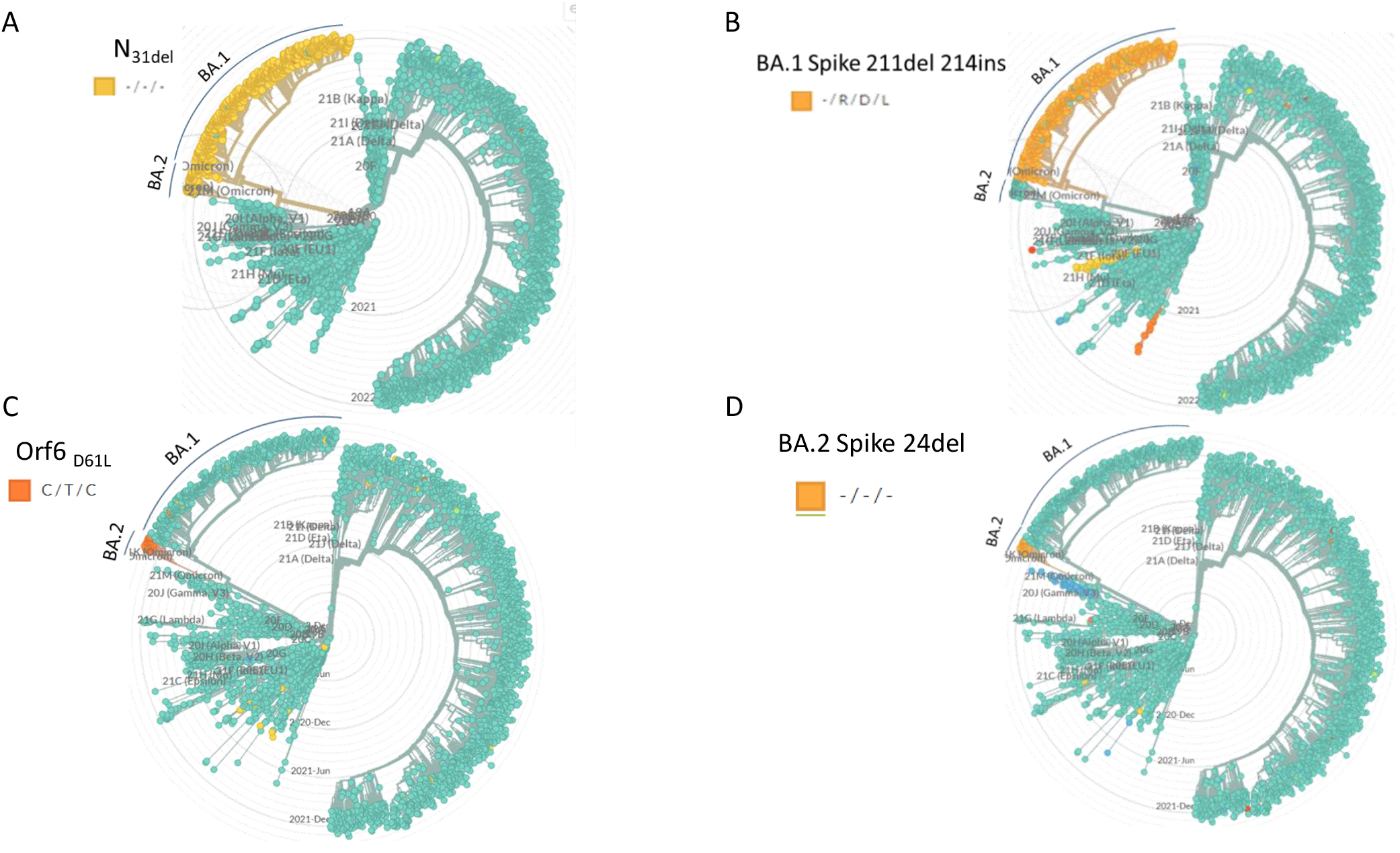
Specificity of selected PCR target mutations. The target mutations selected for qPCR design were identified with respect to circulating strains by using the GISAID-based data with the NextStrain phylogenetic analysis software. The clades that contain the selected mutations are highlighted in each dendrogram. The clade annotation (BA.1, BA.2) is marked with a label and a line.

**Figure 2.**
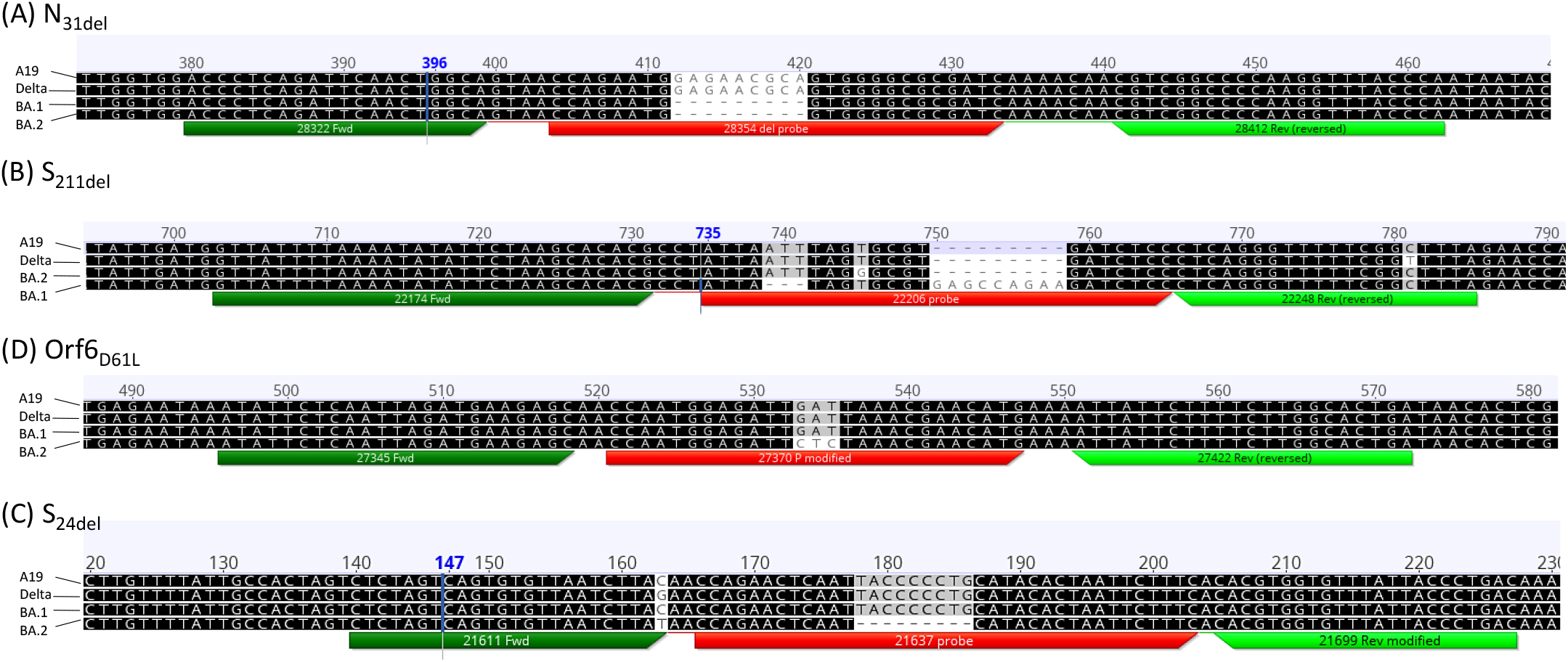
Position of the BA.1/BA.2-specific reaction components. The locations of the primers and probe of each reaction are indicated with respect to the specific mutation. The following sequences were used for the alignments: A19 (Wuhan strain) - NC_045512, Delta sequence: Spain/CT-HUVH-04902/2021|EPI_ISL_2284972, BA.1 sequence: South Africa/NICD-N21672/2021|EPI_ISL_6704871, BA.2 sequence: Israel 8001158. Analysis was performed using the MUSCLE alignment tool embedded in the Geneious software package.

### 3.2. Evaluation of the analytical sensitivity of the new reactions

The S_24del_ reaction was combined in a multiplex that contained also the E-sarbeco inclusive reaction and the endogenous control human RNAse P reaction. A pool of IVT targets corresponding to the multiplex reaction tested, was serially diluted and used with the new multiplex. Three multiplex reactions were tested: E-sarbeco + N_31del_ + S_24del_ + hRNAse P, E-sarbeco + S_211del_, and E-sarbeco + Orf6_D61L_. Based on the calibration curve analyses, the analytic limit of detection for the E-sarbeco, hRNAseP, N_31del_, and S_24del_ was determined to be between1 and 10 copies per reaction (**Figure 3A-D**).

**Figure 3.**
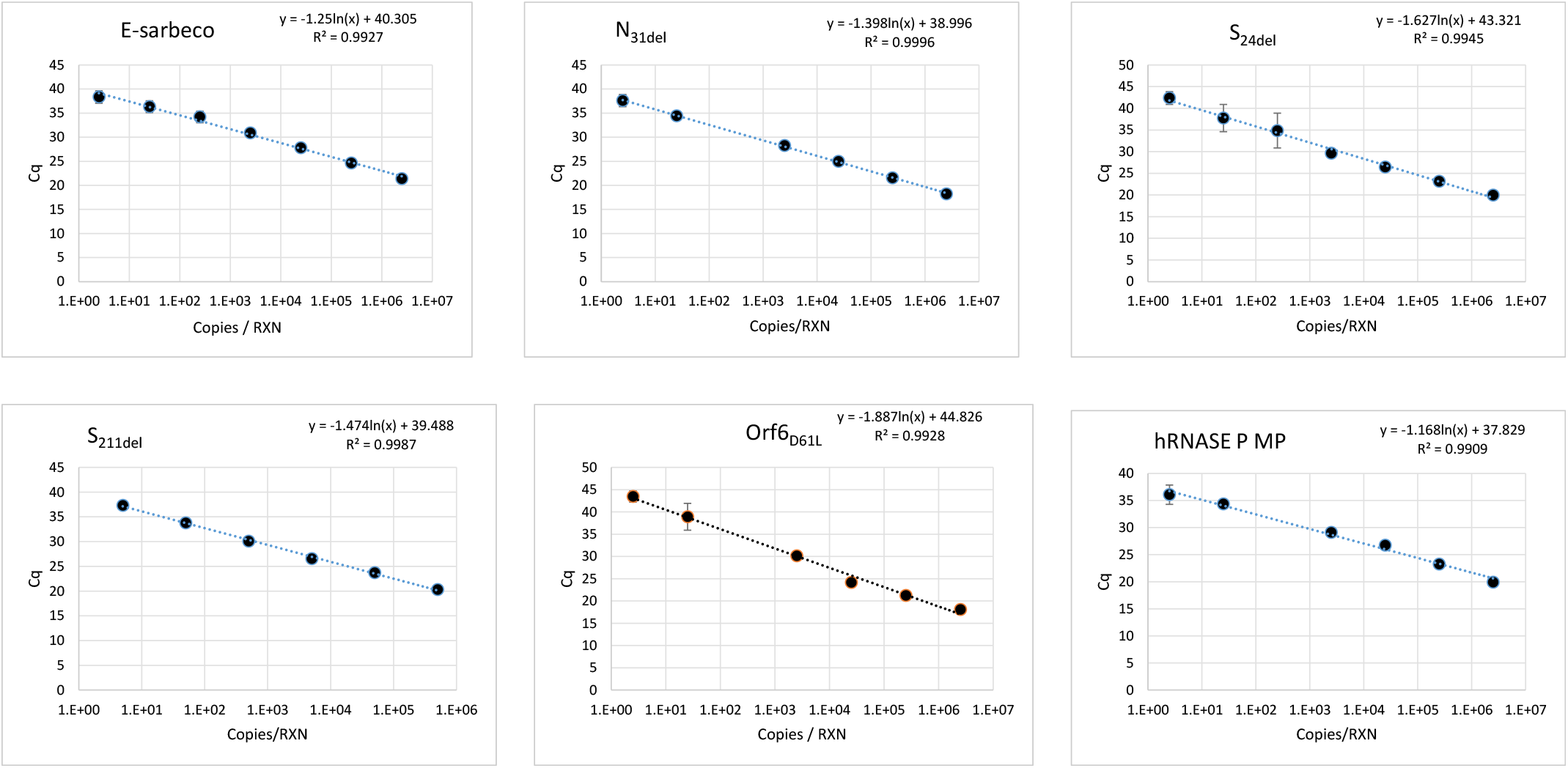
Calibration of the multiplex reactions sensitivity. *In vitro* transcribed RNA molecules containing the target sequence of each reaction were serially diluted and examined using the following reactions: E-sarbeco, N_31del_, S_24del_, S_211del_, Orf6_D61L_ and RNAse P. The Cq values obtained for each dilution in each reaction were plotted against the calculated concentration of target RNA copies. Target copies in concentrations down to 250 copies/reaction (Copies/RXN) were run in 8 repeats each. Target copies in a concentration below 250 Copies /RXN were run in 20 repeats. The R^2^ value and the formula derived from the regression line of each reaction plot are shown.

### 3.3. Identification of BA.1 and BA.2 clinical samples

Clinical samples previously identified as Delta (B.1.617.2), BA.1 and BA.2 by sequencing were tested in different combinations of the inclusive and selective reactions. Out of 36 samples examined, four were identified as Non-BA.1/BA.2 (all were previously identified as Delta), 10 were identified as BA.1 (previously identified as Omicron) and 21 were identified as BA.2 (**Table 3**). One sample (51039) was positive only in the N_31de_l reaction with a Cq value of 38.99 and was therefore classified as “Unclassified sample”. The Cq values of the samples ranged between nine (Cultured sample) and 41 (Weak clinical sample), demonstrating that the sensitivity of the reactions with clinical samples was consistent with the results of the analytical sensitivity calibration.

**Table 3.** Examination of clinical samples using the BA.1 and BA.2 reactions. Each sample was tested using combinations of the inclusive E-sarbeco reaction and the novel BA.1/BA.2 reactions. Nan: No amplification. N.T.: Not tested. The classification of each sample based on the results is indicated.

|  | All COV19 | BA.1+2 | BA.1 ONLY | BA.2 ONLY | BA.2 ONLY | Internal control |  |
| --- | --- | --- | --- | --- | --- | --- | --- |
| Sample | Cov19 E | N-31del | S-211DEL | Orf6-D61L | S-24del | RNAse P | Classification |
| 1796 | 26.75 | 28.12 | NaN | 29.76 | 29.31 | 32.00 | BA.2 |
| 21499 | 31.00 | 33.73 | NaN | 26.02 | 30.10 | 24.00 | BA.2 |
| 23164 | 24.97 | NaN | NaN | NaN | NaN | 28.37 | Non-Omicron |
| 23166 | 23.70 | NaN | NaN | NaN | NaN | 24.93 | Non-Omicron |
| 23172 | 18.84 | NaN | NaN | NaN | NaN | 31.76 | Non-Omicron |
| 23173 | 24.88 | NaN | NaN | NaN | NaN | 25.23 | Non-Omicron |
| 50804 | 16.70 | 15.48 | 17.39 | NaN | NaN | 24.74 | BA.1 |
| 50858 | 15.10 | 13.75 | 13.63 | NaN | NaN | 29.37 | BA.1 |
| 50859 | 15.22 | 13.46 | 13.17 | NaN | NaN | 25.12 | BA.1 |
| 50860 | 12.08 | 12.46 | 9.03 | NaN | NaN | NaN | BA.1 |
| 50862 | 12.40 | 9.73 | 10.56 | NaN | NaN | NaN | BA.1 |
| 50864 | 13.17 | 9.00 | 11.56 | NaN | NaN | NaN | BA.1 |
| 50866 | 12.83 | 10.62 | 10.38 | NaN | NaN | NaN | BA.1 |
| 50867 | 11.50 | 9.26 | 13.07 | NaN | NaN | 26.01 | BA.1 |
| 51030 | 25.65 | 28.82 | 24.98 | NaN | NaN | 29.28 | BA.1 |
| 51038 | 33.50 | 34.33 | 40 | NaN | NaN | 26.63 | BA.1 |
| 51039 | NaN | 38.99 | NaN | NaN | NaN | 30.00 | Unclassified |
| 51040 | 29.98 | 31.70 | NaN | 30.89 | 30.18 | 27.19 | BA.2 |
| 51041 | 21.87 | 24.60 | NaN | 26.11 | 20.82 | 28.90 | BA.2 |
| 51042 | 31.02 | 32.64 | NaN | 27.81 | 29.93 | 30.60 | BA.2 |
| 51044 | 32.47 | 32.85 | NaN | 34.09 | 33.82 | 29.26 | BA.2 |
| 51045 | 34.36 | 35.96 | NaN | 36.65 | 35.06 | 28.44 | BA.2 |
| 51046 | 23.43 | 25.64 | NaN | 29.91 | 24.53 | 27.00 | BA.2 |
| 51048 | 25.57 | 28.08 | NaN | 29.11 | 26.03 | 31.00 | BA.2 |
| 51057 | 39.11 | 40.17 | NaN | 40.76 | 40.50 | 29.72 | BA.2 |
| 51060 | 24.77 | 25.92 | NaN | 27.32 | 26.05 | 29.23 | BA.2 |
| 51062 | 25.96 | 26.83 | NaN | 31.06 | 29.79 | 26.98 | BA.2 |
| 51066 | 19.05 | 21.68 | NaN | 18.76 | 18.76 | 38.00 | BA.2 |
| 51068 | 29.46 | 30.53 | NaN | 31.66 | 32.19 | 27.84 | BA.2 |
| 51070 | 24.92 | 26.17 | NaN | 29.11 | 26.97 | 29.03 | BA.2 |
| 51081 | 23.64 | 25.08 | NaN | 27.40 | 23.03 | 30.62 | BA.2 |
| 51085 | 27.91 | 29.05 | NaN | 30.78 | 30.33 | 32.03 | BA.2 |
| 51094 | 20.13 | 20.83 | NaN | 23.56 | 23.08 | 27.93 | BA.2 |
| 51097 | 33.01 | 33.68 | NaN | 37.71 | 34.84 | 26.09 | BA.2 |
| 51100 | 37.25 | 34.98 | NaN | N.T. | 36.25 | 28.48 | BA.2 |
| 51105 | 36.91 | 35.73 | NaN | N.T. | 26.26 | 30.71 | BA.2 |

## DISCUSSION

The constant need to rapidly identify new and emerging SC-2 variants for clinical, epidemiological and research purposes necessitates vigilant and continuous development of new tools. Since the onset of the SC-2 pandemic, the increased infectivity of the SC-2 variants compared with the initial strain, led to their unprecedented spread in the entire globe, within a matter of a few months (Volz *et al*. 2021, Chakraborty *et al*. 2021, Lambrou *et al*. 2022). Due to their increased infectivity and pathogenicity, the effect of the spreading of different SC-2 variants on public health can be significant and so is the demand for adequate diagnostic tools. While whole-genome sequencing is ultimately the best classification tool, its implementation as a rapid, high-throughput, and available diagnostic tool worldwide, is not yet applicable. Commercial products, such as the Seegene Novaplex and Osang GeneFinder SARS-COV-2 variant kits (https://seegenetech.com, www.osanghc.com) can identify specific mutations, which are associated with major SC-2 variants. However, these kits often target mutations that are not exclusive to a particular variant, such as the spike N501Y or 69-70 deletion, or mutations that can occur, although in a low frequency, independently of the variant type, such as the spike L452R. Furthermore, some of these kits require subsequent testing of the same sample more than once, and the assumed variant identity is determined based on a combination of several separate tests (www.thermofisher.com/). The reactions described herein are specific and can distinguish between BA.1/BA.2 and non-BA.1/BA.2, in a single assay. The ability to distinguish between closely related, but not identical variants may have both clinical and research consequences, and its affordability may render it applicable for immediate use in many diagnostic laboratories, where genomic (or Sanger-based) sequencing is not available. Lastly, positive identification of BA.1 and/or BA.2 RNA is advantageous when testing environmental samples, which usually represent a heterogeneous pool of many individual sources. Negative identification, such as the negative signal in the Thermo SC-2 detection kit, cannot be used in such cases, since the absence of signal can result from low sensitivity, and not necessarily due to the presence of the mutation. However, in the absence of a better tool, the negative identification was initially used to identify the Alpha variant (Davies *et al*. 2021), and is now used by some laboratories to identify suspected BA.1 samples. The new assays described herein are straightforward, sensitive, modular and provide specific and positive identification of BA.1 and BA.2 samples. They may therefore be of immediate use for diagnostic and research laboratories, as a useful tool for detecting and differentiating between the currently circulating BA.1/BA.2 variants.

## Data Availability

All data produced in the present study are available upon reasonable request to the authors

